# Transmission of corona virus disease 2019 during the incubation period may lead to a quarantine loophole

**DOI:** 10.1101/2020.03.06.20031955

**Authors:** Wei Xia, Jiaqiang Liao, Chunhui Li, Yuanyuan Li, Xi Qian, Xiaojie Sun, Hongbo Xu, Gaga Mahai, Xin Zhao, Lisha Shi, Juan Liu, Ling Yu, Meng Wang, Qianqian Wang, Asmagvl Namat, Ying Li, Jingyu Qu, Qi Liu, Xiaofang Lin, Shuting Cao, Shu Huan, Jiying Xiao, Fengyu Ruan, Hanjin Wang, Qing Xu, Xingjuan Ding, Xingjie Fang, Feng Qiu, Jiaolong Ma, Yu Zhang, Aizhen Wang, Yuling Xing, Shunqing Xu

## Abstract

**Background:** The ongoing outbreak of novel corona virus disease 2019 (COVID-19) in Wuhan, China, is arousing international concern. This study evaluated whether and when the infected but asymptomatic cases during the incubation period could infect others.

**Methods:** We collected data on demographic characteristics, exposure history, and symptom onset day of the confirmed cases, which had been announced by the Chinese local authorities. We evaluated the potential of transmission during the incubation period in 50 infection clusters, including 124 cases. All the secondary cases had a history of contact with their first-generation cases prior to symptom onset.

**Results:** The estimated mean incubation period for COVID-19 was 4.9 days (95% confidence interval [CI], 4.4 to 5.4) days, ranging from 0.8 to 11.1 days (2.5th to 97.5th percentile). The observed mean and standard deviation (SD) of serial interval was 4.1±3.3 days, with the 2.5th and 97.5th percentiles at −1 and 13 days. The infectious curve showed that in 73.0% of the secondary cases, their date of getting infected was before symptom onset of the first-generation cases, particularly in the last three days of the incubation period.

**Conclusions:** The results indicated the transmission of COVID-9 occurs among close contacts during the incubation period, which may lead to a quarantine loophole. Strong and effective countermeasures should be implemented to prevent or mitigate asymptomatic transmission during the incubation period in populations at high risk.

## Introduction

The corona virus disease (COVID-19) has spread rapidly throughout China and poses an increasing threat to globe health.^1^ The disease is caused by a novel coronavirus (SARS-CoV-2), which was first discovered in the epidemic center of Wuhan, Hubei province, China in December 2019.^2^ Although China has taken unprecedented actions to halt the outbreak (e.g. lockdown of Wuhan), the reported case numbers are still rising exponentially, particularly in Wuhan. As of February 17, 2020, a total of 72436 COVID-19 cases were officially confirmed in mainland China,^3^ and 42752 of which come from Wuhan.^4^ The rapidly increasing number of cases in Wuhan promotes people to concern whether there are infected cases not identified or quarantined in time.

The prior outbreak of severe acute respiratory syndrome (SARS) and Middle East respiratory syndrome (MERS), which were caused by corona virus, were reported to be rarely transmitted during the asymptomatic incubation period.^5-7^ An urgent question that scientists are racing to learn is whether an asymptomatic COVID-19 case during the incubation period can infect others.^8^ Another major unanswered question is when these asymptomatic cases in the incubation period can infect others, and whether individuals in contact with the COVID-19 cases before the symptom onset should be isolated and provided with medical observation. Until now, only one article reported transmission of SARS-CoV-2 infection from an asymptomatic case in the incubation period in Germany.^9^ One article from Germany found SARS-CoV-2 in a throat swab in two asymptomatic cases,^10^ and the other article from China suggested the transmission by an asymptomatic carrier of SARS-CoV-2 in a familial cluster of five COVID-19 patients.^11^ Compared with the relative small number of asymptomatic cases, the majority of COVID-19 cases when they have no symptoms in their incubation period may contribute to a significant number of infection cases.

All the close contacts with COVID-19 cases are required to be isolated in China.^12^ However, only the close contacts with confirmed patients after symptom onset are put in quarantined in Wuhan city, but other close contactors with the infected cases before the symptom onset are not isolated due to the restrained resources. If the asymptomatic transmission during the incubation period contributes to a significant number of infection cases, a loophole of quarantine will ensue.

In the present study, we aimed to examine whether and when the COVID-19 cases during their incubation period infect others, in order to offer clues to containment measures to mitigate the spread of infection.

## Methods

### Study design and data collection

Accurate estimation of the incubation period is essential to clarify whether the cases who are in the incubation period are a potential source of infection. The exposure time of the majority cases in Wuhan is ambiguous and therefore cannot be used to estimate the incubation period. By examining the cases who were confirmed to be infected with SARS-CoV-2 after a short visit to Wuhan or having a short contact history with a confirmed case, the incubation period could be accurately estimated based on their precise exposure time. Moreover, identification of people in households or communities who had contacted with a confirmed COVID-19 case before symptom onset could help to evaluate the potential transmission during the incubation period. We collected the data of confirmed COVID-19 cases outside Wuhan city and Hubei province in China from local officials in China. The information included demographic characteristics, exposure or contact history, time of exposure and symptom onset, and the relationship of confirmed cases in clusters. Data were entered into a database, and verified by two independent authors. Discrepancies were resolved by discussion and consensus.

In order to increase the accuracy of the incubation period estimation, the selected cases were limited to those with an accurate exposure period of no more than three days. In addition, we only selected the cases who had symptom onset before Chinese Spring Festival (January 25, 2020) to reduce the bias of exposure time, since the custom of visiting relatives and friends in Spring Festival may result to the increase of people’s contact and exposure to unknown infectious source. The cases for estimation of the incubation period were harvested according to the criteria as follows: having a short travel history to Wuhan or having a contact history with a confirmed case, and having no other potential source of infection; and their stay duration or contact exposure was no more than three days, and the information on the dates of their exposure and symptom onset was available. Finally, 106 cases were included in for the analysis of incubation period.

The data of family or community clustered COVID-19 cases throughout February 16, 2020 were used to assess the potential of transmission during the asymptomatic incubation period. We defined the first-generation cases in a cluster as those who had a travel history to Wuhan, or had a contact history (contact with the people from Wuhan, or confirmed cases, or persons with respiratory symptoms), and had been confirmed to have infection of SARS-CoV-2. The secondary cases were defined as those who had a clear contact history to the first-generation cases, and had been confirmed to have infection of SARS-CoV-2, but had no other potential infection source. Cases in the clusters met the criteria as follows: the secondary cases had a close-contact history with their first-generation cases prior to symptom onset, and the information on the contact time and symptom onset time of the first-generation and the secondary cases was complete. A total of 50 first-generation cases and 74 secondary cases in 50 clusters were included for the analysis.

### Statistical Analysis

We described the demographic characteristics and exposures of the cases, and defined the incubation period as the exposure to the transmission source to the onset of symptoms. As each individual’s incubation period was a typically time-to-event, we used a Weibull distribution-based survival analysis model with the extension of Kaplan-Meier estimator to fit the curve of the incubation period for COVID-19 cases.^13,14^ The 95% confidence intervals for the mean and the percentiles of the incubation period were estimated by a bootstrapping method with resampling of 10,000 times.^15^

The serial interval was defined as the time from symptom onset in the first-generation case to symptom onset in the secondary case. The back-projection method was used to estimate the infectious curve for the secondary cases of the clusters to further investigate the possibility of transmission during the incubation period.^16^ Since the symptom onset date of the first-generation cases in different clusters were different, we centralized the symptom onset dates of all first-generation cases into a specific date, and the symptom onset dates of secondary cases were transformed accordingly. After the transformation, the infectious curve would indicate the number of secondary cases emerging before or after the symptom onset dates of the first-generation cases. We defined the back-projection equation as follows:

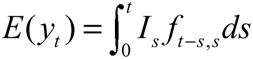

*y*_*t*_ denotes for the number of symptom onset at a transformed time *t*, and *I*_*s*_ denotes the numbers of infected cases at a transformed time *s*, and *f*_*t* − *s, s*_ denotes the probability density function of incubation period for COVID-19, which indicated that the probability of cases infected at a transformed time *t* − *s*, and symptom onset at a transformed time *s*. Using this non-parametric back-projection method, we estimated the infected secondary cases (*It*) at a specific transformed time t.

All analyses were performed in R 3.1 (“surveillance” and “survival” packages) and SAS 9.4.

### Ethics approval

The data of the confirmed cases were from publicly reported, and this study as thus considered exempt from institutional review board approval.

## Results

Of the 106 confirmed COVID-19 cases with a clear travel or contact history, the median age was 41 years (range, 19 to 73), and 70 (66.0%) were known to be male (Table 1). The majority (82.1%) had an exposure period of 1-2 days, and 67.9% had a short travel history to Wuhan city. We estimated the mean incubation period to be 4.9 days (95% confidence interval [CI], 4.4 to 5.4) days, ranging from 0.8 to 11.1 days (2.5th to 97.5th percentile) (Fig. 1).

**Table 1.** Characteristics of the 106 confirmed COVID-19 cases for the estimation of incubation period.

| Characteristics | N (%) |
| --- | --- |
| Median age (range) - yr | 41 (19-73) |
| Age group |  |
| < 18 yr | 0 (0.0) |
| 18-44 yr | 58 (54.7) |
| 45-64 yr | 38 (35.8) |
| ≥ 65 yr | 5 (4.7) |
| Missing | 5 (4.7) |
| Male sex | 70 (66.0) |
| Missing | 3 (2.8) |
| Exposure history<sup>a</sup> |  |
| Wuhan travel history | 72 (67.9) |
| Contact history of confirmed case | 35 (33.0) |
| Exposure period |  |
| 1 day | 57 (53.8) |
| 2 days | 30 (28.3) |
| 3 days | 19 (17.9) |

<sup>a</sup> There was one case who had both a history of travel to Wuhan and contact with a confirmed case.

**Figure 1.**
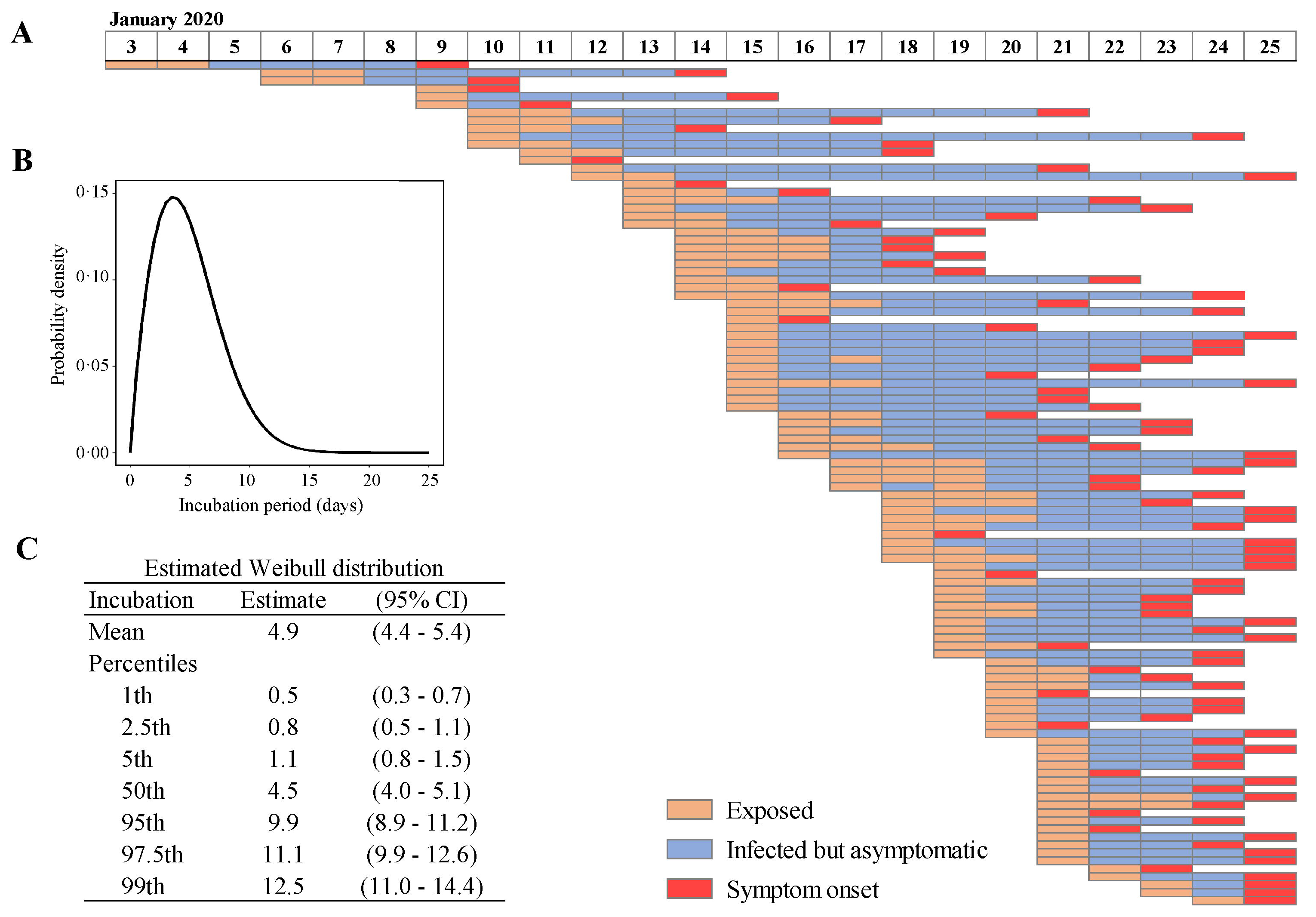
Chorology of exposures and dates of symptom onset of 106 confirmed COVID-19 cases and distribution of the estimated incubation period. (A) Timelines for 106 cases with distinct periods are shown as exposed (orange), infected but asymptomatic (blue), and symptom onset (red). (B) The estimated incubation period distribution. (C) The mean and percentiles of estimated incubation period.

Among the 50 clusters infected with SARS-CoV-2, the median age of the first-generation cases was 47 years (range, 21 to 73), and 58.0% were male, and 82.0% had a travel history to Wuhan (Table 2). The median age of the 74 secondary cases was 45 years (range 7-83), and 45.9% were male. As the timelines shown in Figure 2, most of the secondary cases (89.2%) had symptom onset after their first-generation cases, while four secondary cases (5.4%; in cluster 5, 21, 29, and 46) had symptom onset before their first-generation ones, and four pairs of first-generation and secondary cases (5.4%; in cluster 22, 35, 41, and 43) had symptom onset on the same day. The observed mean and standard deviation (SD) of serial interval was 4.1±3.3 days, with the 2.5th and 97.5th percentiles at −1 and 13 days (Fig. 2).

**Table 2.** Characteristics of the cases in the 50 clusters.

| Characteristics | The first-generation cases<br>(N=50) | The secondary cases<br>(N=74) |
| --- | --- | --- |
| Median age (range) - yr | 47 (21-73) | 45 (7-83) |
| Age group – no./total no. (%) |  |  |
| < 18 yr | 0 (0.0) | 3 (4.1) |
| 18-44 yr | 16 (32.0) | 28 (37.8) |
| 45-64 yr | 20 (40.0) | 12 (16.2) |
| ≥ 65 yr | 6 (16.0) | 20 (27.0) |
| Missing | 8 (16.0) | 11 (14.9) |
| Male sex – no./total no. (%) | 29 (58.0) | 34 (45.9) |
| Missing | 4 (8.0) | 2 (2.7) |
| Exposure history –no./total no. (%)<sup>a</sup> |  |  |
| Wuhan travel history | 41 (82.0) | 0 (0.0) |
| Contact history | 10 (20.0) | 74 (100.0) |

<sup>a</sup>There was one case who had both a history of travel to Wuhan and contact.

**Figure 2.**
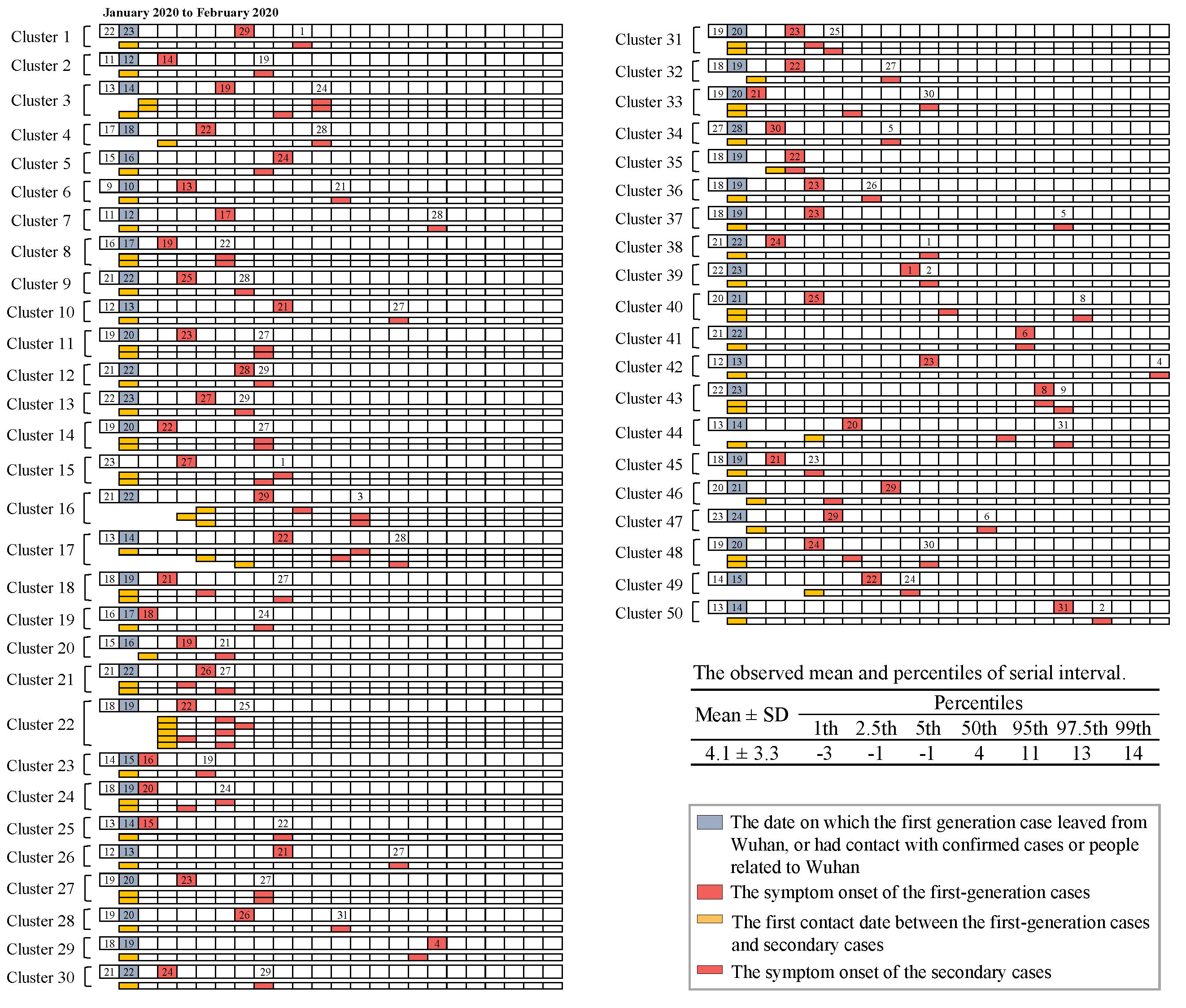
Time lines for 50 clusters and distribution of the observed serial interval. Number in the boxes are calendar dates in January and February 2020.

After the symptom onset dates of the first-generation cases were centralized, the peak of the infectious curve of the secondary cases was shown locating before the symptom onset dates of the first-generation cases (Fig. 3A). The majority of the secondary cases (73.0%) were infected before the symptom onset of the first-generation cases, while 18.9% and 8.1% were infected on and after the date of the symptom onset of the first-generation cases, respectively (Fig. 3B). Particularly, 66.2% of the secondary cases were infected within three days before the symptom onset dates of the first-generation cases.

**Figure 3.**
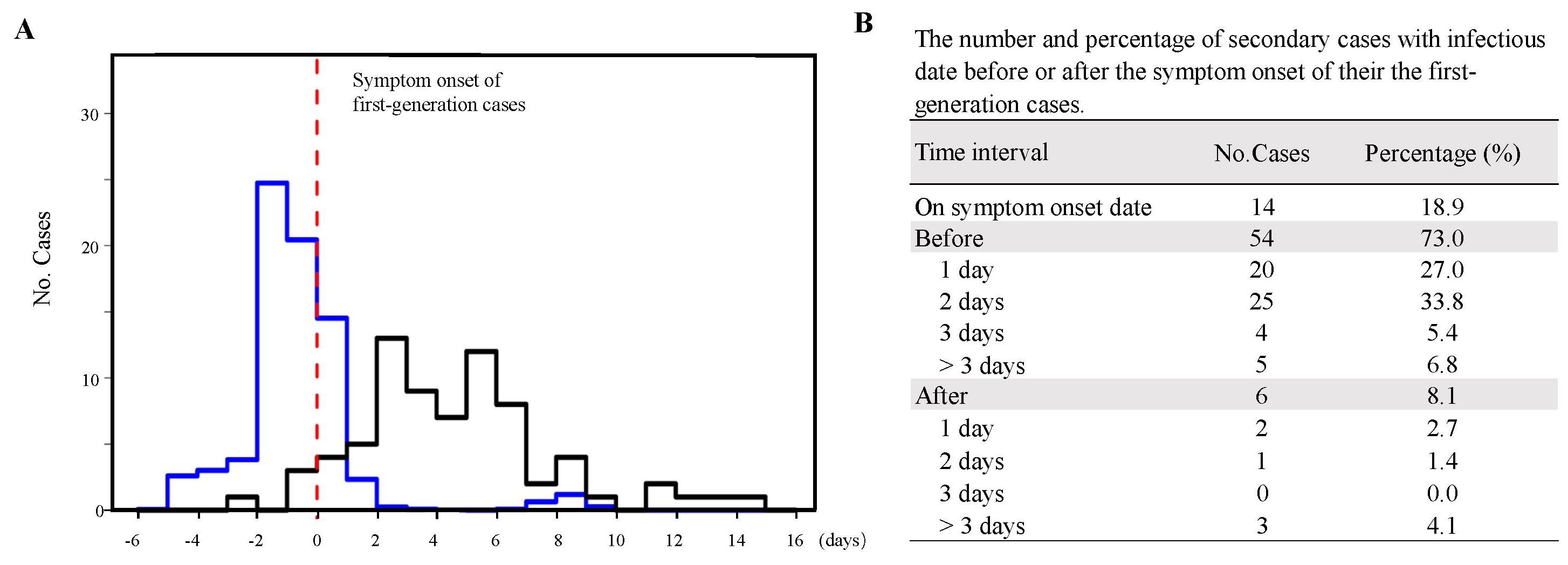
The infectious curve of the secondary cases. (A) The infectious and symptom onset curves for the secondary cases after the symptom onset dates of the first-generation cases were centralized. The curves are shown as the number of the secondary cases on the infected date (blue) and actual symptom onset date (black). The red dotted line indicates the centralized date of symptom onset of the first-generation cases. (B) The number and percentage of the secondary cases with infectious date before or after the symptom onset of their first-generation cases.

## Discussion

Our study initially demonstrated asymptomatic transmission of COVID-19 in the incubation period, especially in the last three days of incubation period, by estimating the incubation period with the use of accurate exposure history of confirmed cases. Among the clusters, we found that four secondary cases had symptom onset earlier than their first-generation cases, which further provides the evidence of the transmission during the incubation period. Our findings could help the officials to take strong and effective measures to contain the spread of COVID-19 and to the prediction of the epidemic trend.

We estimated the mean incubation period of COVID-19 infections to be 4.9 days, which was calculated based on a big sample size with valid data. Most importantly, the exposure time interval of the 106 cases selected in our study was no more than three days, and 82.1% of the cases had an exposure time of 1-2 days, which enabled us to obtain a more accurate estimates of incubation period for COVID-19 than the existing evidence. Li et al. ^17^ reported that the incubation period of the new coronavirus was 5.2 days (95% CI, 4.1 to 7.0) with the 95th percentile at 12.5 days according to the early cases with illness onset exposed to the Huanan seafood wholesale market. The authors deemed that this estimation was somewhat imprecise because it was based on the information of only 10 cases.^17^ Backer et al.^14^ and Lauer et al.^18^ estimated the incubation period based on 34 and 101 publicly reported confirmed COVID-19 cases outside Wuhan. The duration of exposure time of their included cases ranged from one day to more than two weeks, and more than 75% of the cases in the two studies had the exposure time more than three days, which might induce bias in the estimates and cause the estimated incubation period longer than the actual length. In addition, our estimates of incubation period were consistent with the results reported by Guan et al.^19^ that they estimated the mean incubation period less than five days from the data of 1099 confirmed patients in mainland China.

We observed the mean serial interval was 4.1 days for COVID-19, which is shorter than the 8.4-day and the 7.6-day mean serial interval reported for SARS and MERS, respectively.^5,20^ More importantly, the serial interval is shorter than the estimates of the mean incubation period (4.9 days), suggesting that a proportion of secondary transmission may occur before symptom onset of the first-generation cases. Li et al.^17^ estimated the mean serial interval at 7.5 days based on the cases reported in Wuhan city early in the epidemic (mostly in December 2019). However, there were only six pairs first-generation and secondary cases in this study, and the small sampling bias may have been introduced to the variance and mean. Our observed serial interval has been consistently demonstrated by a recent study that the estimated mean serial interval was 4.2 days based on 20 pairs of first-generation and secondary cases.^21^

Our results of infectious curves of the secondary cases in the clusters found that the COVID-19 cases before they feel sick could infect others, and the findings offer some of the best clues to whether individuals in contact with the COVID-19 cases within at least three days prior to the symptom onset should be isolated. Our results indicated that the current quarantine measures have loopholes, which may lead to some asymptomatic cases during the incubation period slipping through the net. At present, the Chinese government has enforced to quarantine all the close contacts of the confirmed cases.^12^ In the Chinese New coronavirus pneumonia prevention and control program (4th ed.), the close contacts who are requried to put in quanrantine are defined as “the individulas who have a history of living together, staying in the same room, or riding in the same vehicles with the suspected cases, clinical diagnosis cases (only in Hubei Province), confirmed cases, and asymptomatic infected persons after they are tested virus positive”.^12^ In this program, only the close contacts after the case has symptom onset or virus positive testing are required to be quarantine, but the close contacts before the case has symptom onset or virus positive testing are not included. If asymptomatic people who are in the incubation period have the potential to spread the virus, the current quarantine measures are inadequate to contain the spread of COVID-19, especially for those individuals and their family members who were isolated together at home.

Due to the shortage of personals for epidemic contain in Wuhan, the close contacts with the confirmed cases prior to symptom onset were not isolated. The quarantine loophole may also explain the occurrence of a lot of familial cluster cases and the rapidly increased number of confirmed cases during the past week in Wuhan city and Hubei province. According to the recent epidemiological study based on a total of 72314 COVID-19 patient records in mainland China, asymptomatic cases accounted for 1.2%.^22^ Compared with the relative small number of asymptomatic cases, the majority of COVID-19 cases when they have no symptoms in their incubation period may contribute to a significant number of infection cases. Therefore, we believe more effective measures should be taken to stem the transmission of asymptomatic contact during the incubation period. Once a case is confirmed, it is imperative to trace back all the close contacts within at least three days prior to the symptom onset, and then quarantine these close contacts. If a patient has extensive social activities before the symptom onset, the risk of transmission will be greatly increased. Therefore, restricting social activities and restricting travel in and out of the cities in severe epidemic areas may effectively reduce the transmission.

All the information about the cases in this study comes from the data released by local authorities, and the cases were screened for analysis by using the same criteria. The information of the cases released by local authorities outside of Hubei province, including the exposure history, was relatively complete and reliable, because less cases were reported outside of Hubei province and staffs working at local center of disease control (CDC) staffs had enough time to interview the cases as soon as possible. Therefore, this study provides a reliable incubation period and a strong evidence of the COVID-19 transmission during the incubation period.

Nevertheless, there are some limitations in this study. The data about virus detection were not available for those asymptomatic persons in their incubation period. Since the follow-up quantitative reverse-transcriptase-polymerase-chain-reaction (qRT-PCR) of virus nucleic acid assay was not available for most close contacts currently in China, it is difficult to obtain evidence of molecular biological monitoring in a short time. If qRT-PCR assays are performed for a group of close contacts, more convincing evidence will be provided for transmission of COVID-19 during the incubation period.

In conclusion, the COVID-19 cases in the incubation period are potential infection sources, especially within three days prior to the symptom onset. It is imperative to trace back the close contacts of confirmed cases before the symptom onset and avoid the quarantine loopholes.

## Data Availability

Data are publicly available.All the information about the cases is released by local authorities in China.

## Contributors

SX had the idea for and designed the study and had full access to all data in the study and take responsibility for the integrity of the data and the accuracy of the data analysis. WX, JL, CL, and YL contributed to writing of the report and the statistical analysis. SX contributed to critical revision of the report. CL, XQ, XS, HX, YL, GM, YX, XZ, LS, JL, MW, QW, AN, YL, JQ, QL, XL, SC, SH, JX, FR, HW, QX, XD, XF, FQ, JM, YZ, AW, and YX contributed to collecting data, creating of the tables and figures and the statistical analysis All authors contributed to data acquisition, data analysis, or data interpretation, and reviewed and approved the final version.

## Declaration of interests

All authors declare no competing interests.

## Acknowledgement

We are very grateful to Liu Liu and Chaohong Yu in the Editorial Department of Current Medical Science, Liuyu Ivy Chen in founder of TransWords.net, Henry M. Clever, Ph.D. student in the healthcare robotics lab at Georgia Institute of Technology, and Yanjian Wan in Wuhan Centers for Disease Prevention & Control for editing our manuscript.

## Notes

### Competing Interest Statement

The authors have declared no competing interest.

### Funding Statement

No external funding was received.

